# COVID-19 vaccine effectiveness against progression to in-hospital mortality — Zambia, 2021-2022

**DOI:** 10.1101/2022.07.18.22277749

**Authors:** Duncan Chanda, Jonas Z Hines, Megumi Itoh, Sombo Fwoloshi, Peter A Minchella, Khozya D. Zyambo, Suilanji Sivile, Davies Kampamba, Bob Chirwa, Raphael Chanda, Katongo Mutengo, Mazinga F. Kayembe, Webster Chewe, Peter Chipimo, Aggrey Mweemba, Simon Agolory, Lloyd B. Mulenga

## Abstract

**Background:** COVID-19 vaccines are highly effective for reducing severe disease and mortality. However, vaccine effectiveness data is limited from sub-Saharan Africa, where SARS-CoV-2 epidemiology has differed from other regions. We report COVID-19 vaccine effectiveness against progression to in-hospital mortality in Zambia.

**Methods:** We conducted a retrospective cohort study among admitted patients at eight COVID-19 treatment centers across Zambia during April 2021 through March 2022. Patient demographic and clinical information including vaccination status and hospitalization outcome (discharged or died) were collected. Multivariable logistic regression was used to assess the odds of in-hospital mortality by vaccination status, adjusted for age, sex, number of comorbid conditions, disease severity, and COVID-19 treatment center. Vaccine effectiveness of ≥1 vaccine dose was calculated from the adjusted odds ratio.

**Results:** Among 1,653 patients with data on their vaccination status and hospitalization outcome, 365 (22.1%) died. Overall, 236 (14.3%) patients had received ≥1 vaccine dose before hospital admission. For patients who had received ≥1 vaccine dose, 22 (9.3%) died compared with 343 (24.2%) among unvaccinated patients (p <0.01). The median time since receipt of a first vaccine dose was 52.5 days (IQR: 28-107). Vaccine effectiveness for progression to in-hospital mortality among hospitalized patients was 64.8% (95% CI: 42.3-79.4%).

**Conclusions:** Among patients admitted to COVID-19 treatment centers in Zambia, COVID-19 vaccination was associated with lower progression to in-hospital mortality. These data are consistent with evidence from other countries demonstrating benefit of COVID-19 vaccination against severe complications. Vaccination is a critical tool for reducing the consequences of COVID-19 in Zambia.

**Key points:**

- Receipt of ≥1 COVID-19 vaccine dose reduced progression to in-hospital mortality in Zambia by 64.8%
- Mortality benefit of COVID-19 vaccines was sustained during the period of omicron transmission in Zambia

## Introduction

Since the onset of the COVID-19 pandemic, >170,000 persons have died of COVID-19 in sub-Saharan Africa (1). In Zambia, from March 20, 2020 to July 13, 2022, a total of 327,570 confirmed cases and 4,009 confirmed deaths were recorded (2), although the true number of infections and deaths was likely greater (3,4).

As of February 2022, African countries had received more than 587 million vaccine doses, enough to vaccinate ∼360 million people (which is ∼37% doses needed to reach vaccination targets) (1). However, through April 2022, only 14% of the eligible population in sub-Saharan Africa had been fully vaccinated (1). The ChAdOx1-S COVID-19 vaccine became available in Zambia in April 2021 and approximately one year later, ∼2.3 million persons in Zambia were fully vaccinated (22% of persons aged ≥12 years, the eligible population) (2). In addition to poor access to COVID-19 vaccines for much of 2021, several other challenges hindered vaccination programs in Zambia, including sub-optimal access to preventive health care/routine immunization, under-resourced health care systems, inadequate cold chain systems, lack of awareness of benefits of vaccination, and vaccine misconceptions and misinformation.

COVID-19 vaccines have demonstrated excellent efficacy in clinical trials and good effectiveness in real-world observational studies, including against variants of concern and in people who have had prior COVID-19 infections (5-7). In particular, COVID-19 vaccines substantially reduce risk of severe disease and mortality (8,9). Yet, there is paucity of vaccine efficacy and effectiveness data from countries in sub-Saharan Africa. Most of the vaccine evidence from Africa have originated from South Africa, where several vaccine clinical trials have been conducted and observational vaccine effectiveness (VE) studies have been published (5,10-13). A single dose of the Ad26.COV2.S vaccine was associated with lower hospitalizations and mortality during beta and delta waves in South Africa (11), and additional evidence demonstrated effectiveness against COVID-19 hospitalizations during the omicron wave (5). In Zambia, full receipt of a primary COVID-19 vaccine was associated with lower SARS-CoV-2 infections and symptomatic illness during an outbreak in a prison when omicron was the dominant variant (13). However, data from other countries in sub-Saharan Africa is limited to date.

Filling the COVID-19 vaccine evidence gap in Africa is important to counter misinformation and skepticism toward research from other parts of the world (14,15). Furthermore, SARS-CoV-2 epidemiology in Africa has differed from elsewhere, with fewer confirmed cases and deaths, lower symptomatic rates, yet high evidence of infection-induced immunity (16-18). Furthermore, many countries in sub-Saharan Africa have a high burden of persons living with HIV (PLHIV) and tuberculosis, which are conditions associated with increased risk of poor outcomes from COVID-19 (19,20). Lastly, differences in social and health system structures might impact vaccine delivery. Therefore, more real-world data on COVID-19 vaccines from countries in Africa are needed. We assessed COVID-19 VE against progression to in-hospital mortality in patients hospitalized with COVID-19 in Zambia.

## Methodology

We conducted a retrospective cohort study of patients admitted to COVID-19 treatment centers in Zambia to assess COVID-19 VE against progression to in-hospital mortality between April 15, 2021 (when Zambia first began offering COVID-19 vaccines) and March 31, 2022. Since the pandemic onset, patients diagnosed with COVID-19 requiring in-patient admission in Zambia were admitted to COVID-19 treatment centers that had specifically designated isolation and treatment units staffed by clinicians and nurses trained in COVID-19 clinical management (21). The COVID-19 clinical outcomes study was conceived during the early weeks of the COVID-19 epidemic in Zambia and eventually included eight COVID-19 treatment centers in five cities: Lusaka (four treatment centers), Ndola, Kitwe, Kabwe, and Livingstone (22). Data on relative variant genome frequency by region from GISAID (Global Initiative on Sharing Avian Influenza Data) were utilized to define the SARS-CoV-2 variant that was circulating during each wave (23). ChAdOx1-S COVID-19 vaccine was the first vaccine type available in Zambia and by late 2021, Ad26.COV2.S, mRNA-127, BNT162b2, and Sinopharm BBIBP-CorV were available in the country. An additional vaccine dose after completing a primary vaccine series (i.e., “booster”) became available in Zambia in January 2022. The study protocol was approved by the University of Zambia Biomedical Research Ethics Committee; it was also reviewed in accordance with the US Centers for Disease Control and Prevention (CDC) human research protection procedures and was determined to be research, but CDC investigators did not interact with human subjects or have access to identifiable data or specimens for research purposes.

Patients who provided verbal consent for clinical care at participating treatment centers had demographic and clinical data collected at admission and during hospitalization until they were discharged or died. Data were retrospectively abstracted from clinical records using a standardized case record form adapted from WHO (24) and entered into REDCap (Research Electronic Data Capture) electronic data capture tools by trained staff at a later date (25). Vaccination information was collected from patients during clinical care and recorded in their charts. Proof of vaccination included self-report and/or review of vaccine card. For those with missing vaccine information, attempts were made to cross-reference the electronic national vaccine registry to improve data completeness. Severe COVID-19 was defined as having an oxygen saturation (SpO2) <90%, respiratory rate >30 breaths/minute, or need for oxygen therapy (26). The number of self-reported comorbidities was summed for each patient, including cardiac disease, hypertension, diabetes, other pulmonary disease, HIV, tuberculosis (active or previous), asthma, kidney disease, liver disease, neurological disorders, asplenia, malignant neoplasms, and current smoking. History of prior COVID-19 was not available for patients, although the reliability of this history is not clear given a low case detection proportion in Zambia (3).

Full vaccination was defined as receiving the 1st dose of a one-dose vaccine or 2nd dose of a two-dose vaccine ≥14 days prior to COVID-19 treatment center admission. Partial vaccination was defined as receiving the 1^st^ dose of a two-dose vaccine ≥14 days prior to admission but either not yet receiving the 2nd dose or receiving the 2nd dose ≤13 days prior to admission. Those vaccinated with their 1^st^ COVID-19 vaccine dose ≤13 days prior to admission were considered indeterminate and those with missing vaccine dates were classified as such. Because vaccination dates were missing for many patients with known vaccination status, we also created a separate category of patients who received ≥1 COVID-19 vaccine dose and used this as our primary predictor variable (with full/partial vaccination status being secondary analyses). The main outcome variable was categorized as discharged from hospital or died in the hospital. Length of stay was calculated as the days between admission and discharge or death. Patients whose vaccination status or hospitalization outcomes could not be determined were excluded from the analyses.

The analysis was restricted to patients admitted during the period that COVID-19 vaccines were available in Zambia (April 15, 2021, through March 31, 2022). We used chi-square test and student’s t-test to compare categorical and continuous variables, respectively. We used multivariable logistic regression to calculate the odds of in-hospital mortality by vaccination status, adjusting for age, sex, number of comorbid conditions, disease severity at admission, hospitalization month, and COVID-19 treatment center. VE was calculated as 1 minus the adjusted odds ratio times 100. We did secondary analyses to calculate VE by full/partial vaccination status, vaccine type, the predominant circulating variant (i.e., delta from April to November 2021, and omicron [subvariants B1 and B2 (23)] from

December 2021 through March 2022 (27)), and HIV status. Additionally, we analyzed if there was an association between progression to in-hospital mortality and predominant variant periods (delta versus omicron waves), adjusting for patients’ vaccination status.

## Results

Overall, 2,385 persons had data on their hospitalization course abstracted at a COVID-19 treatment center in Zambia during April 2021 through March 2022. Of patients with data abstracted, 1,821 (76.4%) had vaccination status documented (815 [61.5%] persons during the delta variant predominant period and 1,006 [94.9%] persons during the omicron variant predominant period). Among patients with known vaccination status, 1,653 (69.3% of total) had a known hospitalization outcome (Figure 1).

**Figure 1.**
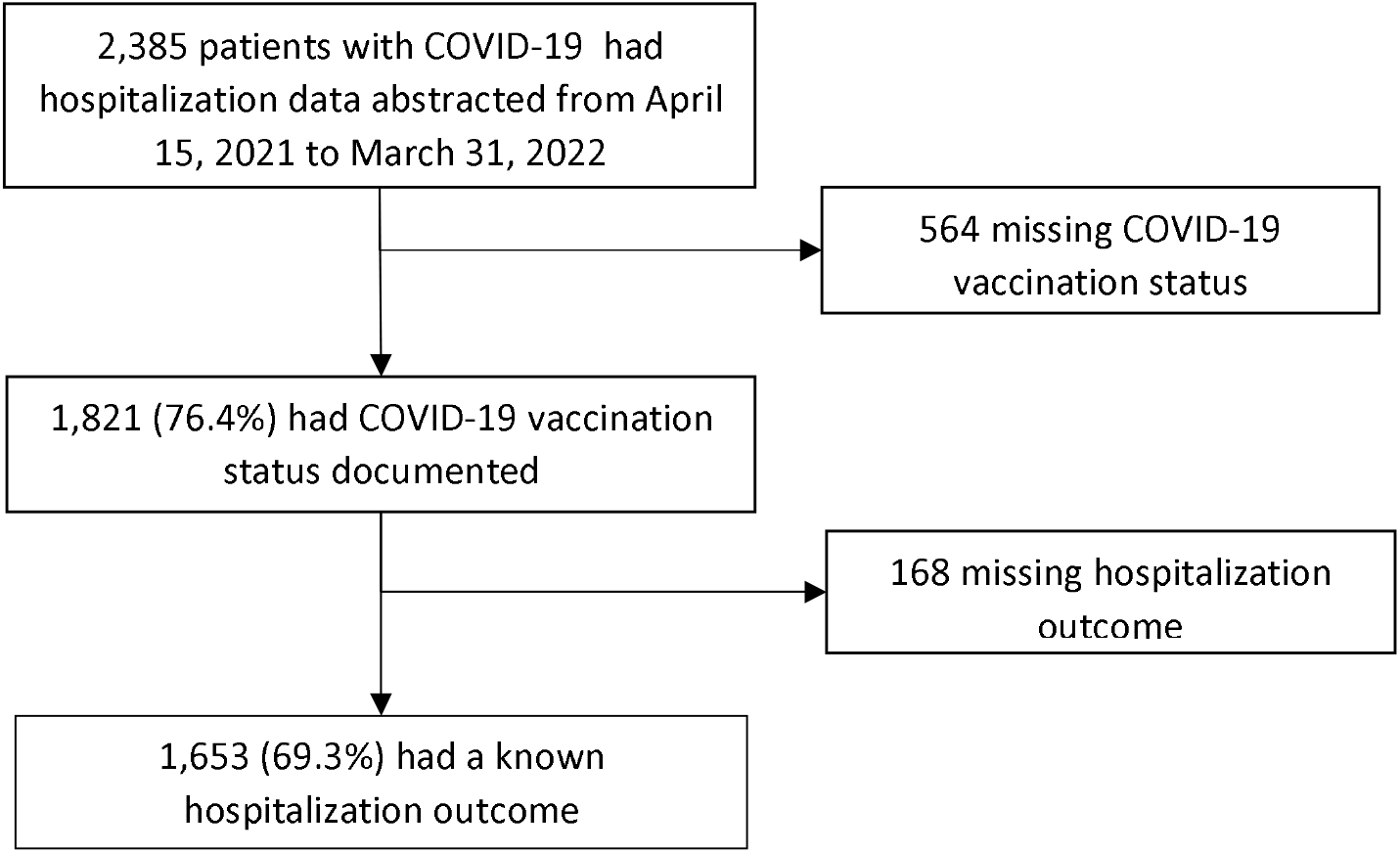
Sample size flow diagram for analysis of COVID-19 vaccine effectiveness against in-hospital mortality in Zambia, 2021-2022

The median age of participants was 47 years (interquartile range [IQR]: 30-65 years) and females accounted for 852 (51.5%) patients (Table 1). Overall, 1,065 (64.4%) patients reported having at least one comorbidity, with 266 (17.9%) patients reporting having HIV infection (compared to national prevalence of 11.1% in persons aged 15-49 years (28)). The average length of stay was 2.5 days and 365 (22.1%) patients died during their hospitalization.

**Table 1.**
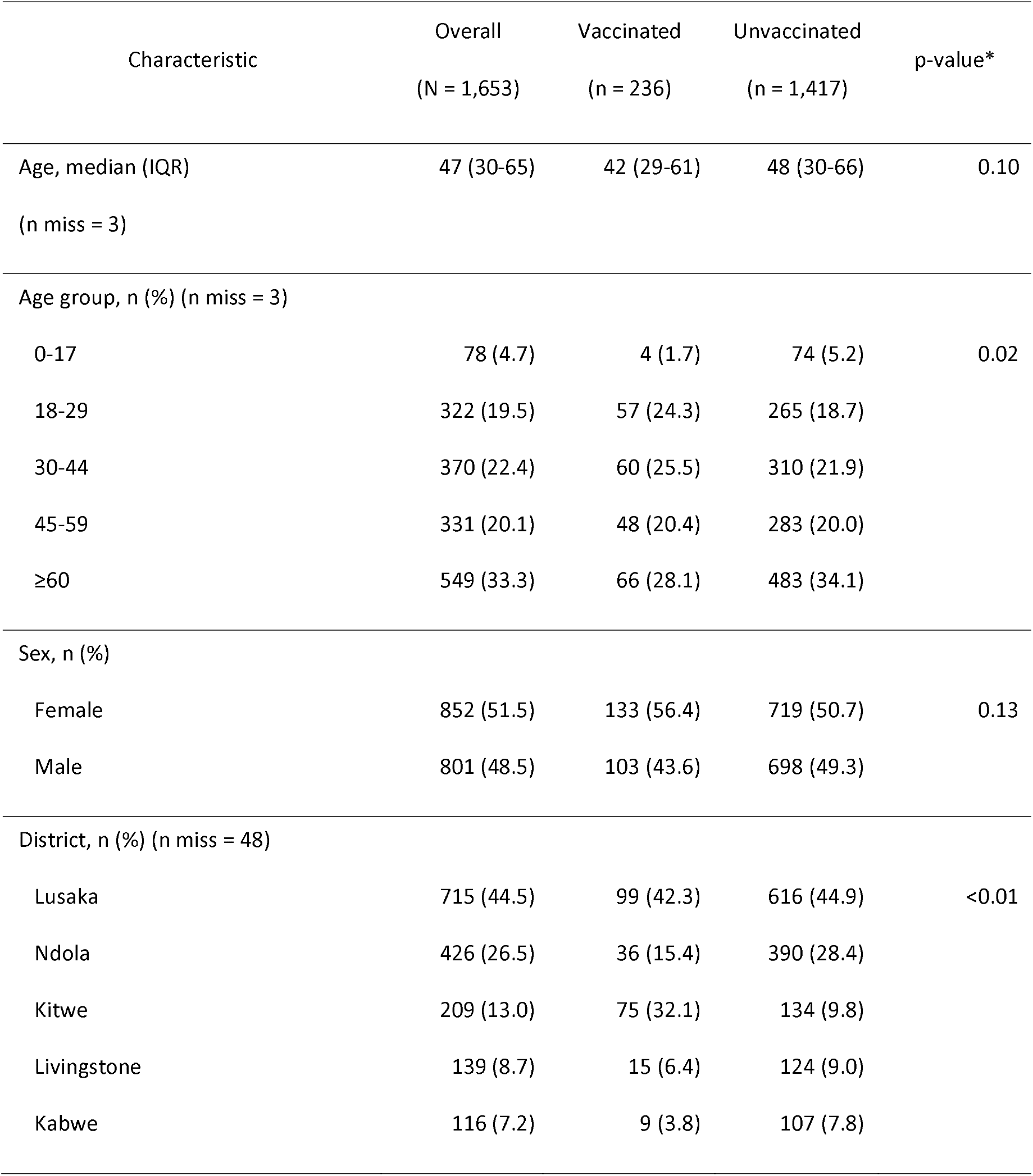
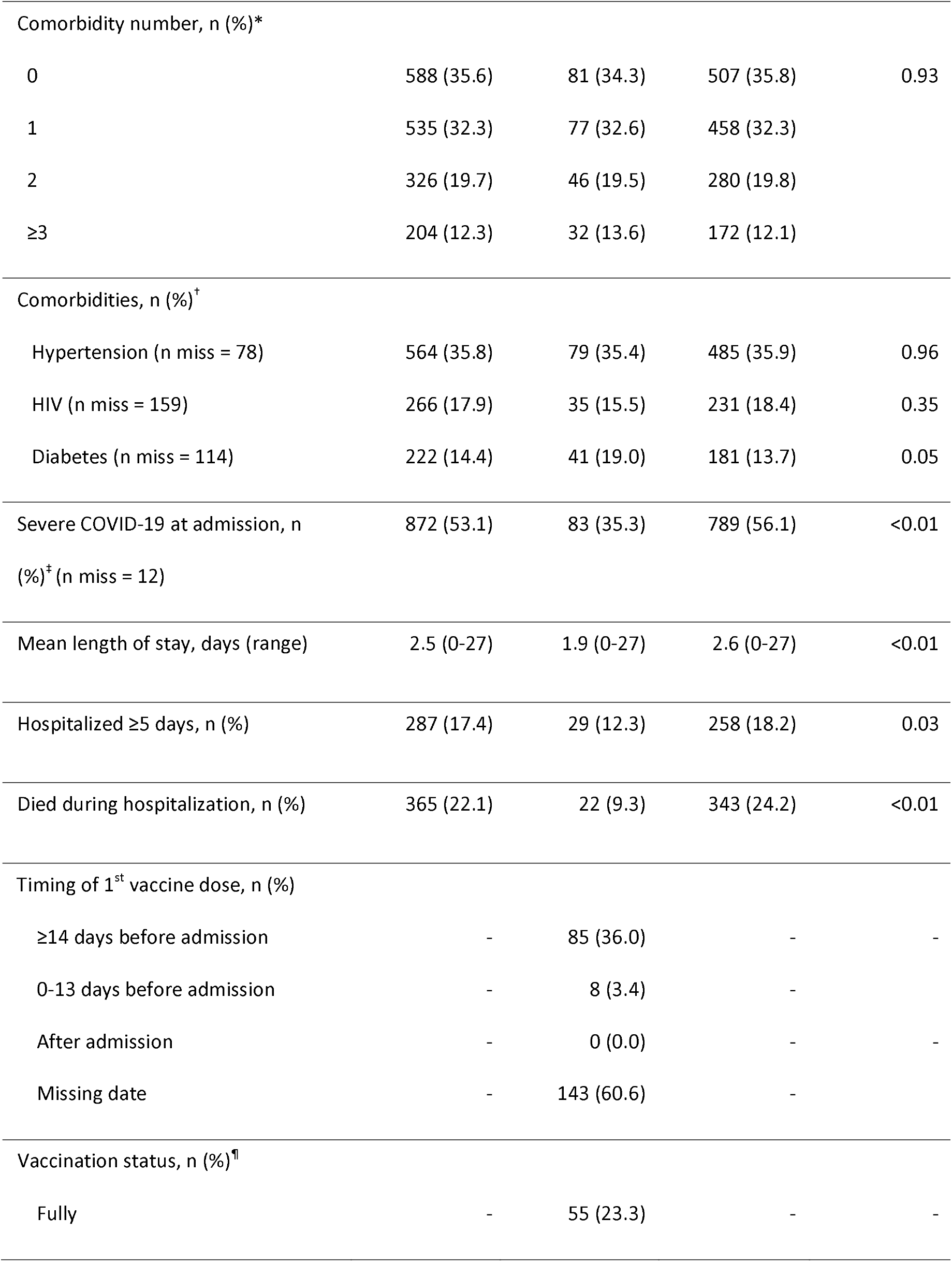
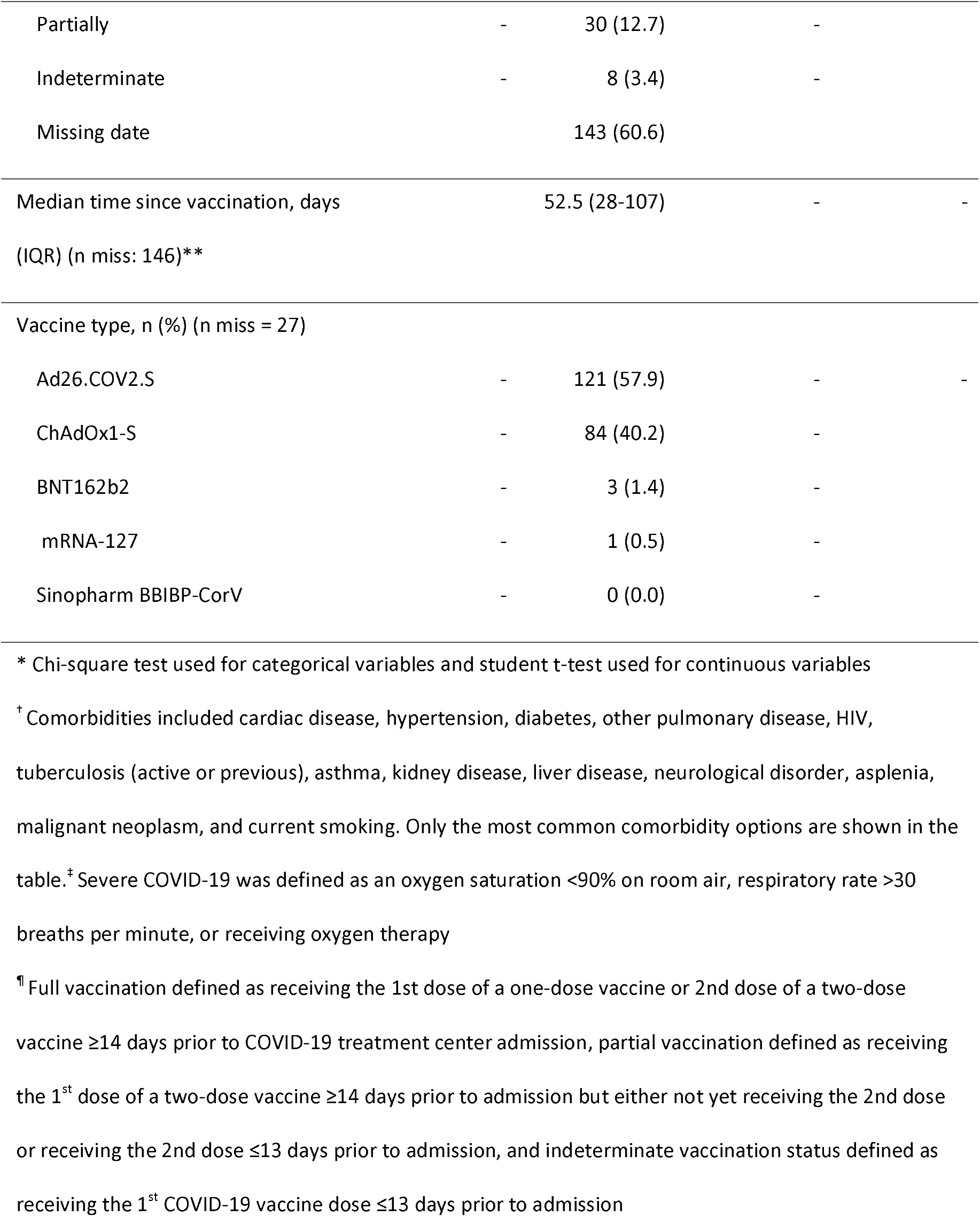
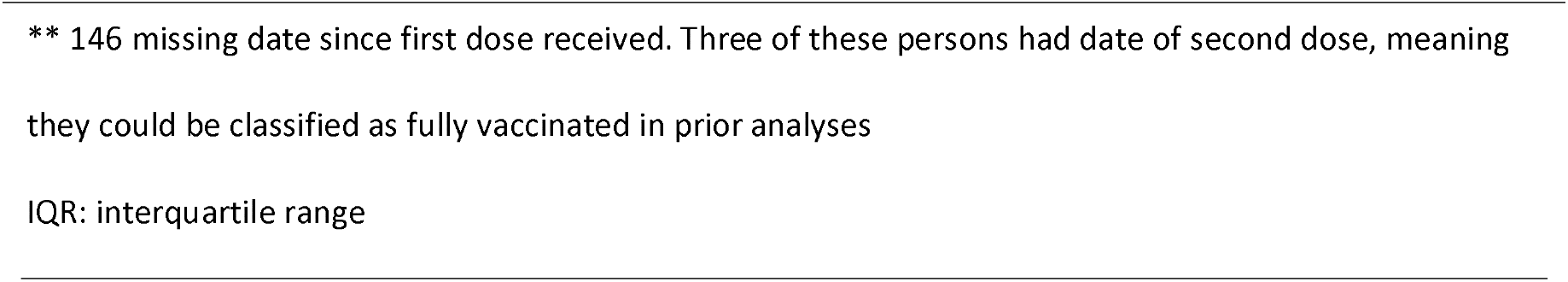
Characteristics and outcomes of patients admitted to COVID-19 treatment centers with known hospitalization outcome in Zambia, 2021-2022

Two-hundred thirty-six (14.3%) patients had received ≥1 vaccine dose before hospital admission, of whom 55 (23.3%) were fully vaccinated, 30 (12.7%) were partially vaccinated, 8 (3.4%) had an indeterminate vaccination status, and 143 (60.6%) were missing information on vaccination date(s) (Table 1). No patient had a record of receiving a 3^rd^ vaccine dose. The median time since receipt of a first vaccine dose was 52.5 days (IQR: 28-107).

The average length of stay for patients with ≥1 vaccine dose was 1.9 days versus 2.6 days for unvaccinated patients (p <0.01,) (Table 1). Twenty-two (9.3%) patients reporting ≥1 vaccine dose died compared to 343 (24.2%) unvaccinated patients (p <0.01). VE of ≥1 COVID-19 vaccine dose against progression to in-hospital mortality was 64.8% (95% confidence interval [CI]: 42.3-79.4) (Table 2). VE of ≥1 vaccine dose during the period when delta was dominant was 65.0% (95% CI: 22.5-85.8) and during the period when omicron was dominant was 64.8% (95% CI: 31.4-82.9).

**Table 2.** COVID-19 vaccine effectiveness against progression to in-hospital mortality - Zambia, April 2021-March 2022

| Vaccination status | Deaths in vaccinated group*, n (%) | Vaccine effectiveness, % (95% CI) |  |
| --- | --- | --- | --- |
|  |  | Crude VE | Adjusted VE<sup>†</sup> |
| ≥1 vaccine dose (n = 236) | 22 (9.3) | 67.8 (50.4-80.1) | 64.8 (42.3-79.4) |
| Vaccination status<sup>‡</sup> |  |  |  |
| Full (n = 55) | 6 (10.9) | 61.7 (16.5-85.4) | 60.9 (2.3-86.5) |
| Partial (n = 30) | 4 (13.3) | 51.8 (-24.7-85.9) | 61.3 (-15.8-89.7) |
| Indeterminate (n = 8) | 0 (0.0) | - | - |
| Missing (n = 143) | 12 (8.4) | 71.3 (49.7-85.1) | 63.4 (29.7-82.3) |
| Time period (predominant variant)<sup>¶, **</sup> |  |  |  |
| April to November 2021 (delta) (n = 46) | 8 (17.4) | 55.3 (7.5-80.9) | 65.0 (22.5-85.8) |
| December 2021 to March 2022 (omicron) (n = 190) | 14 (7.4) | 59.6 (30.2-78.3) | 64.8 (31.4-82.9) |
| Vaccine type<sup>¶</sup> |  |  |  |
| ChAdOx1-S (n = 84) | 9 (10.7) | 62.1 (32.9-80.5) | 65.2 (33.5-83.1) |
| Ad26.COV2.S (n = 121) | 10 (8.3) | 69.9 (48.2-84.0) | 61.1 (26.6-80.8) |

\* 343 (24.2%) patients died in the unvaccinated group (n = 1,417)
<sup>†</sup> Adjusted for age, sex, number of comorbid conditions, disease severity at admission (defined as oxygen saturation <90% on room air, respiratory rate >30 breaths per minute, or receiving oxygen therapy), hospitalization month, and COVID-19 treatment center
<sup>‡</sup> Full vaccination defined as receiving the 1st dose of a one-dose vaccine or 2nd dose of a two-dose vaccine ≥14 days prior to COVID-19 treatment center admission, partial vaccination defined as receiving the 1<sup>st</sup> dose of a two-dose vaccine ≥14 days prior to admission but either not yet receiving the 2nd dose or receiving the 2nd dose ≤13 days prior to admission, and indeterminate vaccination status defined as receiving the 1<sup>st</sup> COVID-19 vaccine dose ≤13 days prior to admission
<sup>¶</sup> Estimate for receipt of ≥1 COVID-19 vaccine dose
\*\* In the delta variant predominant period, there were 706 unvaccinated patients, of whom 226 (32.0%) died during hospitalization. In the omicron variant predominant period, there were 711 unvaccinated patients, of whom 117 (16.5%) died during hospitalization.
CI: confidence interval; VE: vaccine effectiveness

Among the 266 patients with HIV, 5 (14.3%) who had received ≥1 COVID-19 vaccine dose died compared to 61 (26.4%) who were unvaccinated (p = 0.18). VE of ≥1 COVID-19 vaccine dose against progression to in-hospital mortality among PLHIV was 53.6% (−15.6-84.7).

In-hospital mortality among patients in the study was higher during the delta variant predominant period than the omicron variant predominant period (31.1% vs. 14.5%; p<0.01; odds ratio: 2.7 [95% CI: 2.1-3.4]) (Table 3). Adjusting for the different vaccination coverage of patients between waves (6.1% during delta versus 21.1% during omicron) did not appreciably change the odds of in-hospital mortality (adjusted odds ratio: 2.4 [95% CI: 1.9-3.1]).

**Table 3.**
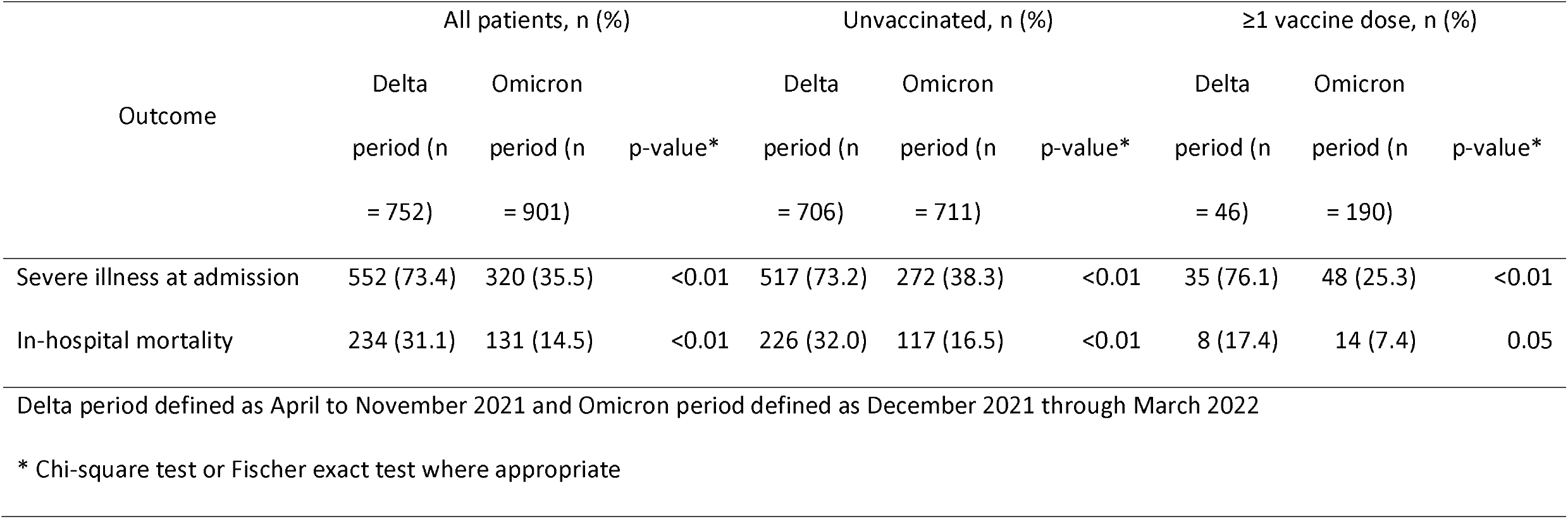
In-hospital mortality by COVID-19 pandemic period (proxy for predominant variant) and vaccination status — Zambia, 2021-2022

| Outcome | All patients, n (%) |  |  | Unvaccinated, n (%) |  |  | ≥1 vaccine dose, n (%) |  |  |
| --- | --- | --- | --- | --- | --- | --- | --- | --- | --- |
|  | Delta | Omicron | p-value* | Delta | Omicron | p-value* | Delta | Omicron | p-value* |
|  | period (n | period (n |  | period (n | period (n |  | period (n | period (n |  |
|  | = 752) | = 901) |  | = 706) | = 711) |  | = 46) | = 190) |  |
| Severe illness at admission | 552 (73.4) | 320 (35.5) | <0.01 | 517 (73.2) | 272 (38.3) | <0.01 | 35 (76.1) | 48 (25.3) | <0.01 |
| In-hospital mortality | 234 (31.1) | 131 (14.5) | <0.01 | 226 (32.0) | 117 (16.5) | <0.01 | 8 (17.4) | 14 (7.4) | 0.05 |

Delta period defined as April to November 2021 and Omicron period defined as December 2021 through March 2022
\* Chi-square test or Fischer exact test where appropriate

## Discussion

Patients vaccinated against COVID-19 had lower progression to in-hospital mortality in Zambia, a finding that was consistent during periods when delta and omicron were the dominant variants circulating in the country. These data from Zambia expand on limited evidence of COVID-19 VE in Africa (5,10-13). COVID-19 vaccination uptake in Africa has lagged other regions but evidence suggests that they are a critical tool for reducing morbidity and mortality from COVID-19. Sharing local evidence of the benefit of COVID-19 vaccine in Zambia might increase vaccine uptake.

Omicron variant of SARS-CoV-2 has mutations that enable it to evade prior immunity, resulting in decreased VE (6). However, protection from severe COVID-19 outcomes is the most conserved vaccine effect across different variants, especially following a booster dose (9,29,30). In our study, there was no decrement in VE against progression to in-hospital mortality when omicron was the predominant variant in Zambia (compared to when delta was predominant). Patients in this analysis had been vaccinated relatively recent (i.e., less than two months), which could explain preserved VE (6); additionally, data from before the omicron surge in the U.S. indicate preserved VE against hospitalization of a single dose adenovirus vaccine for at least six months (31,32).

Even though VE remained similar between the delta and omicron waves in this study, the in-hospital mortality risk was lower during the omicron wave. However, this mortality difference was not driven by higher vaccination coverage among patients during the omicron wave. This finding is similar to those in a study by Modes et al. demonstrating that in one U.S. hospital, fewer fully vaccinated patients died in-hospital during the omicron wave compared to the delta wave (33). This suggests other factors like patient characteristics (e.g., greater proportion of patients with infection-induced immunity during the omicron wave, higher risk patients admitted during the delta wave), system/environmental factors (e.g., relative availability of hospital beds or oxygen supply in Zambia, which were much more constrained during the delta wave than during the omicron wave), and/or virus characteristics (e.g., omicron lower replication competence in lung parenchyma (34)) could be responsible for the mortality difference by wave in Zambia (20).

Some clinical trials for COVID-19 vaccines included PLHIV, although the numbers were too small to derive definitive evidence of efficacy in PLHIV (12,35). Based on immunological studies, vaccines are expected to work well in this high-risk population (36). The Ad26.COV2.S vaccine has been demonstrated to reduce hospitalizations and deaths among PLHIV in South Africa (11), and in Russia, the Gam-COVID-VAC vaccines was shown to be effective in PLHIV, especially for severe disease (37). Although COVID-19 vaccination among PLHIV appeared to be effective against progression to in-hospital mortality in this study, the effect was not statistically significant. The inclusion of additional PLHIV in this study could provide a clearer evidence on this important topic in Zambia, where there is a high burden of HIV (28).

Vaccinated patients in this study were also less likely than unvaccinated patients to be admitted to treatment centers with severe COVID-19, suggesting that vaccination also helped prevent this outcome. Although vaccine coverage in this study was low, it varied by geographic location, suggesting opportunities to learn from best practices in some areas to help other areas increase vaccine coverage.

Our study had several limitations. Data were not available describing either national COVID-19-related hospitalization numbers or COVID-19 hospitalization rates by facility, limiting our understanding of the study’s generalizability. As with many studies conducted during emergency responses, data completeness for some key variables was a challenge that impacted our sample size and resulted in wide confidence intervals. Additionally, the lack of vaccination dose numbers and dates for many participants necessitate use of a non-standard category (i.e., ≥1 vaccine dose) for the primary analysis to avoid excessive loss of participants; the similarity of VE estimates for full and missing vaccination statuses could indicate that many patients with missing vaccination status were in fact fully vaccinated. Additionally, given high levels of infection-induced immunity in Africa (16), hybrid immunity likely confers substantial protection against severe consequence of COVID-19 in Zambia (38). Variant periods were defined based on predominant variants in GISAID data in Zambia and not on sequencing results from patients in the study. HIV status was self-reported, which could have led to biased VE findings in PLHIV. Lastly, there are potentially other unaccounted for confounders which couldn’t be factored into the VE analysis.

Vaccination is a critical tool to reduce the consequences of the SARS-CoV-2 epidemic. These findings are among the first from countries in Africa, and such real-world evidence of COVID-19 vaccines could help increase vaccination in a region that lags the rest of the world by providing local evidence of vaccine benefits in preventing COVID-19 mortality. Substantially increasing COVID-19 vaccinations will help Zambia reach targets and reduce COVID-19 mortality in the country.

## Data Availability

All data produced in the present study are available upon reasonable request to the authors

## Acknowledgements

Lydia Chama (University Teaching Hospital, Lusaka); Martha Ilunga (Kawama Health Center, Kabwe); Florence Chanda Miti (Levy Mwanawasa University Teaching Hospital, Lusaka); Evelyn Mwamba (University Teaching Hospital, Lusaka); Naomi Charity Mwananyau (Levy Mwanawasa University Teaching Hospital, Lusaka); Francis D. Mwansa (Ministry of Health, Lusaka); Linda Musonda (University Teaching Hospital, Lusaka); Agness Mutaja (National Heart Hospital, Lusaka); Charles Mutemba (University Teaching Hospital, Lusaka); Prosperllina Ndhlovu (Ndola Teaching Hospital, Ndola); Amideos Nshikita (Maina Soko Medical Centre, Lusaka); Christabel Phiri (Levy Mwanawasa University Teaching Hospital, Lusaka); Constance Phiri (University Teaching Hospital, Lusaka); Miracle Phiri (Levy Mwanawasa University Teaching Hospital, Lusaka); Mary M. Siamupa (Livingstone Central Hospital, Livingstone); Vanessa Tayali (Kabwe Central Hospital, Kabwe); Harriet Nandi Zulu (Ndola Teaching Hospital, Ndola);

## Attribution of Support

This work has been supported by the Zambia Ministry of Health and the U.S. President’s Emergency Plan for AIDS Relief (PEPFAR) through the U.S. Centers for Disease Control and Prevention (CDC) and the CDC Emergency Response to the COVID-19 pandemic.

## Authorship Disclaimer

The findings and conclusions in this report are those of the author(s) and do not necessarily represent the official position of the funding agencies.

## Notes

### Competing Interest Statement

The authors have declared no competing interest.

### Author Declarations

The study protocol was approved by the University of Zambia Biomedical Research Ethics Committee; it was also reviewed in accordance with the US Centers for Disease Control and Prevention (CDC) human research protection procedures and was determined to be research, but CDC investigators did not interact with human subjects or have access to identifiable data or specimens for research purposes.

